# Direct Health Care Costs Associated with Asthma Hospitalizations Before and During the Covid-19 Pandemic in the United States: A Nationwide Inpatient Sample Analysis

**DOI:** 10.1101/2025.01.09.25320273

**Authors:** N. Solanki, B. F. Fakhry, P. Zhang, A. Attaway, J. Zein, W. Tsuang

## Abstract

**Background:** Asthma, a chronic inflammatory airway disorder, can increase the risk of hospitalizations in individuals with viral infections such as COVID-19. The impact of the COVID-19 pandemic on asthma-related hospitalizations in the United States remains unknown.

**Objective:** We hypothesized that the COVID-19 pandemic led to an increase in economic burden to society and a decrease in hospitalization rates for asthma.

**Methods:** We analyzed weighted data from National Inpatient Sample (NIS) between January 1, 2018, and December 31, 2020. The outcomes were asthma hospitalization rates, length of stay (LOS), in-hospital mortality rates, and hospital admission costs.

**Results:** More people were admitted with a primary diagnosis of asthma in 2018 and 2019 compared to 2020 (hospitalization rate per 100,000: 2018: 38.6 versus (vs) 2019:37.0 vs 2020: 21.4; P <0.001). Hospital costs increased (2018: median [IQR] 5,251 [ 3,426, 8,278]; 2019:5,677 [3,725, 8.969]; 2020: 5,881 [3,920-9,216]; P <0.001). Additionally, in-patient mortality rates slightly increased in 2020, rising to 0.44%, compared to 0.20% in 2018 and 0.30% in 2019 (P <0.001). When comparing geographic divisions, the mid-Atlantic division had the highest hospitalization rates, and the Pacific division had the highest hospitalization costs from 2018-2020 (P <0.05).

**Conclusion:** Asthma hospitalizations decreased in 2020 compared to previous years. Those admitted during the pandemic had slightly higher mortality and significantly increased hospital costs. This investigation provides valuable insights for policy makers about shifts in healthcare utilization during the pandemic.

## Introduction

Asthma is a chronic airway disease marked by variable airflow limitation and airway inflammation^1^. The impact of asthma is large in the United States (US), affecting approximately 25 million individuals and placing a substantial burden on healthcare utilization^2^. In 2019, uncontrolled asthma resulted in nearly 1.8 million emergency department (ED) visits,169,330 hospitalizations, and 4.9 million clinic visits in the US^3,4^. Asthma places a considerable economic burden on patients and society with an estimated cost of more than $80 billion between 2008 and 2013 ^5^.

The COVID-19 pandemic disrupted the US healthcare system with widespread social distancing measures. These restrictions resulted in the cancellation of clinics and procedures, thereby reducing patient access to healthcare services^6^. Simultaneously, hospitals faced challenges in managing the influx of acutely ill patients with COVID-19^7^. Despite the high prevalence of COVID-19 infections, both hospitalizations and ED visits for acute asthma cases decreased in several countries^8–12^. According to the National Syndromic Surveillance Program, EDs in the US had a 31% relative decrease in weekly visits for pediatric asthma in 2020 compared to 2019^13^. The United Kingdom demonstrated a similar decrease in visits to the ED for asthma and a decline in hospital admissions from 2019 to 2020^12,14^.

It remains unclear how the pandemic influenced healthcare access, costs, and outcomes for asthma patients. Existing studies primarily focus on the decrease in ED visits and hospital admissions, but they do not comprehensively address the economic impact or changes in healthcare utilization patterns^7,1510,16,17^. We utilized the National Inpatient Sample (NIS) Database, the largest publicly available all-payer inpatient healthcare database, to investigate regional and national estimates of inpatient utilization, access, cost, and outcomes. We hypothesized that the COVID-19 pandemic was associated with a decrease in hospital admissions for patients with asthma along with an increase in hospitalization costs because of longer hospital stays.

## Methods

### Data source

We used the Healthcare Cost and Utilization Project (HCUP)-National Inpatient Sample (NIS) for the years 2018-2020. NIS captures more than 95% of US hospital admissions with nearly 8 million hospitalizations annually. NIS represents a wide array of hospitals sampled to reflect about one-fifth of US community hospitals^18,19^.

### Study design and population

We queried the NIS database between January 1st, 2018, to December 31st, 2020, using the International Classification of Diseases, 10th Revision, Clinical Modification (ICD-10-CM). Patients with a primary diagnosis of asthma (ICD-10-CM code: J45.XX) were included in the study (n = 88,484 patients) (Figure E1). Participants with missing data were excluded.

We performed analysis on patients admitted in 2020 with a primary diagnosis of COVID-19 (ICD-10 codes U07.9, J12.89, B97.29) and a secondary diagnosis of asthma.

The data from NIS are de-identified, thus exempting review by the Institutional Review Board of the Cleveland Clinic.

### Time frame definitions

We organized our data into strata based on years and quarters with Q1 representing January to March, Q2 for April to June, Q3 for July to September, and Q4 for October to December (Figure E2). We defined the period spanning from January 1, 2018, to December 31, 2019 as the pre-COVID-19 era. The timeframe from January 1, 2020 to December 31, 2020 represented the early COVID-19 era.

### Outcome measures and variables

The primary outcome was hospital admission cost. The secondary outcome variables included asthma hospitalization rate, length of stay (LOS), and in-hospital mortality rate. The HCUP’s NIS provides data on total charges for each hospital, reflecting the billed amount for all services rendered. Cost was adjusted for inflation according to consumer price index (CPU) for medical services in December 2020 (Appendix 1)^20^. The LOS was estimated in days after excluding those who died in the hospital. We categorized LOS into two groups LOS≥ 7 days and LOS <7 days.

We compared outcomes between pre-COVID-19 and the early COVID-19 periods. In addition, we examined disparities in outcomes across age groups, sex, race, household income, and insurance type (Table E1, E2 and Appendix 1-3). Outcome variables were stratified based on geographic division determined by the US Census Bureau^21^. The states representing each of the nine Census Divisions are shown in Figure E3.

### Statistical analysis

To generate national estimates, we used analytic sample weights (discharge-level weight: DISCWT) for patient level observations as described by HCUP guidelines ^22^. We represented categorical variable as percentages, and unadjusted comparisons between groups and were performed using Chi-squared test. Continuous and non-symmetrical variables were represented as medians [interquartile range], and differences were tested using the Mann–Whitney U test/Wilcoxon rank-sum test.

We conducted weighted regression analysis. Multivariate logistic regression was used for categorical outcomes (in-patient mortality, LOS> 7days), and presented as adjusted odds ratios with 95% confidence intervals (CI) (Appendix 3). In all multivariate models, we included all parameters described in Table E1 and tested for multicollinearity. All tests were two-tailed, and the level of statistical significance was set at P <0.05. All analyses were performed using JMP software (Cary, NC, USA).

## Results

### Annual Cost of Asthma Hospitalizations

Overall median asthma hospitalization cost was 5,552 [3,635, 8,751] USD. It rose from 5,251 [3,426, 8,278] USD in 2018 to 5,881 [3,920, 9,216] USD in 2020 (Table 1, Figure E4). The second quarter of 2020 had the lowest cost related to asthma hospitalization during the pandemic (Figure 1).

**Figure 1:**
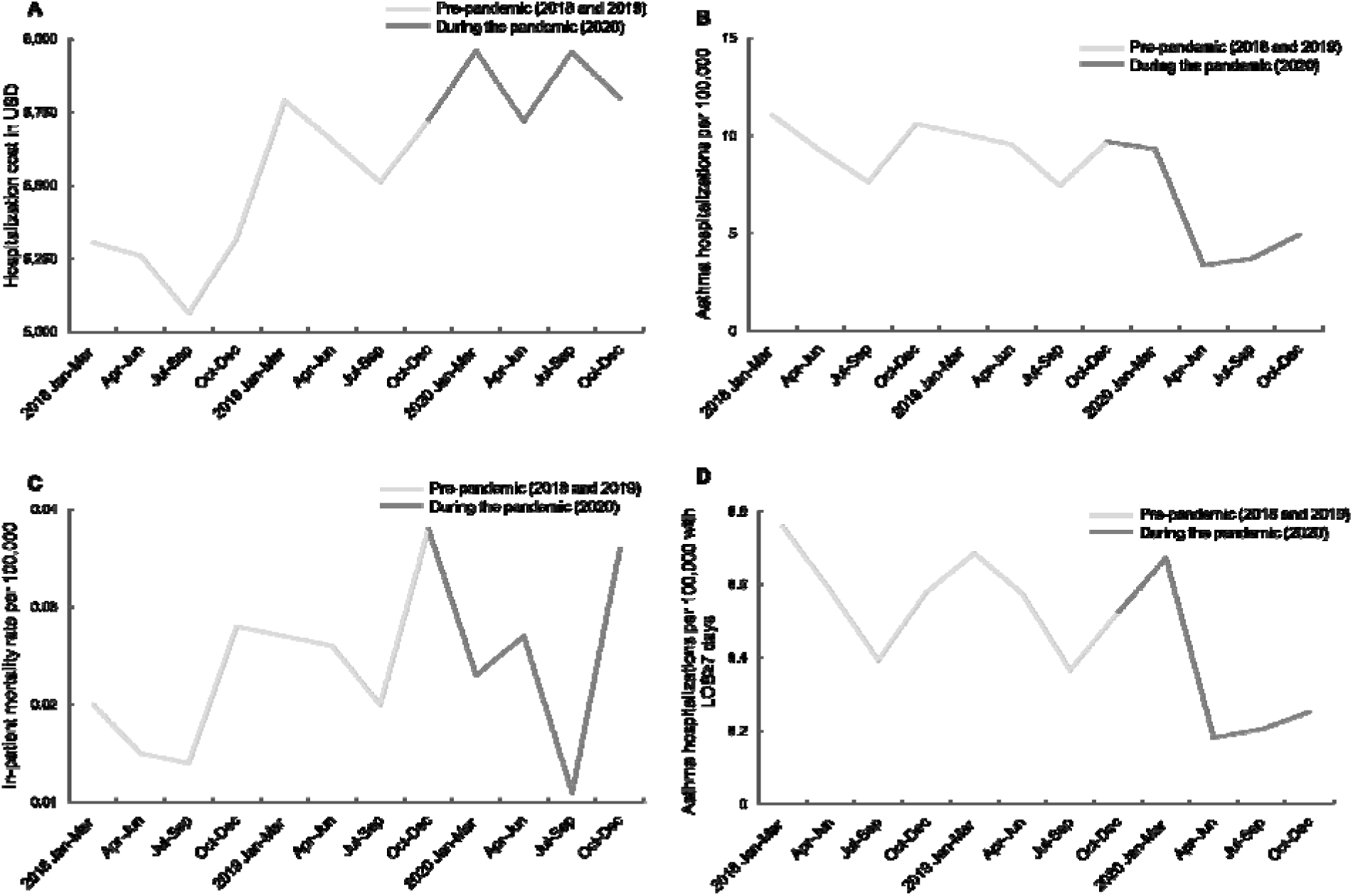
Quarterly changes in healthcare utilization patterns and patient outcomes. A- Hospitalization cost. B- Asthma hospitalizations per 100,000 people. C- In-patient mortality rate per 100,000 people. D-Asthma hospitalizations per 100,000 people with LOS≥ 7 days.

**Table 1.**
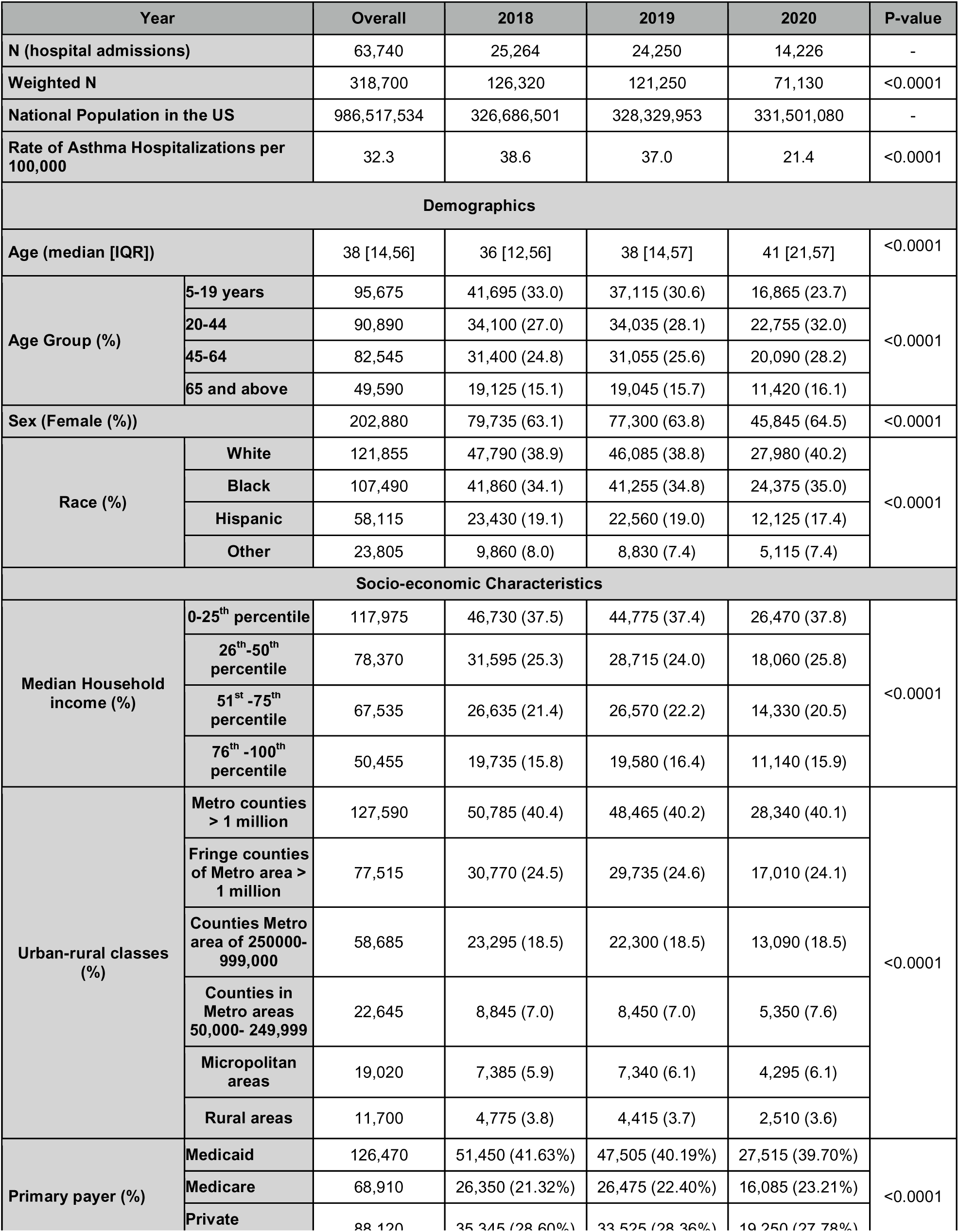
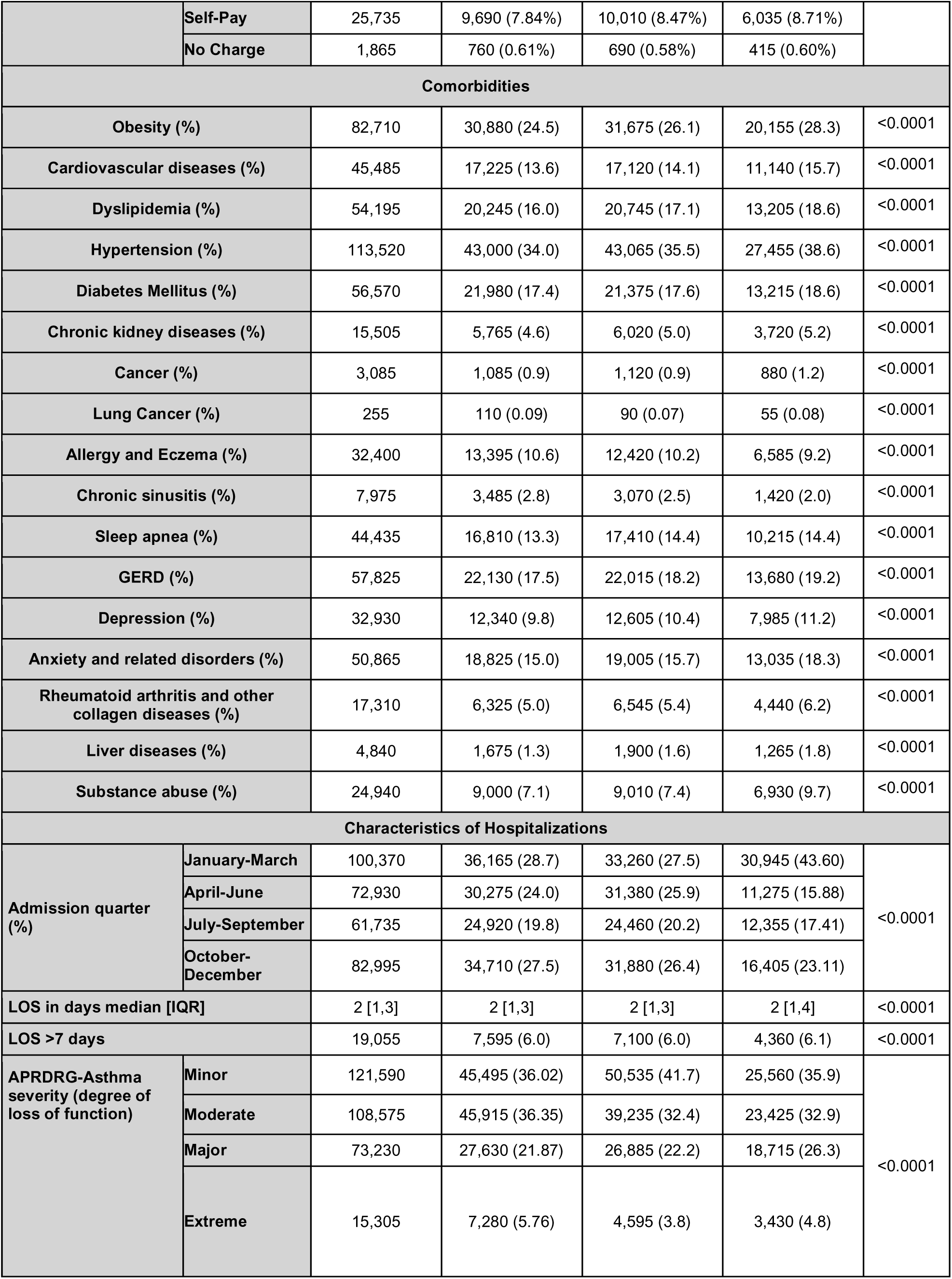
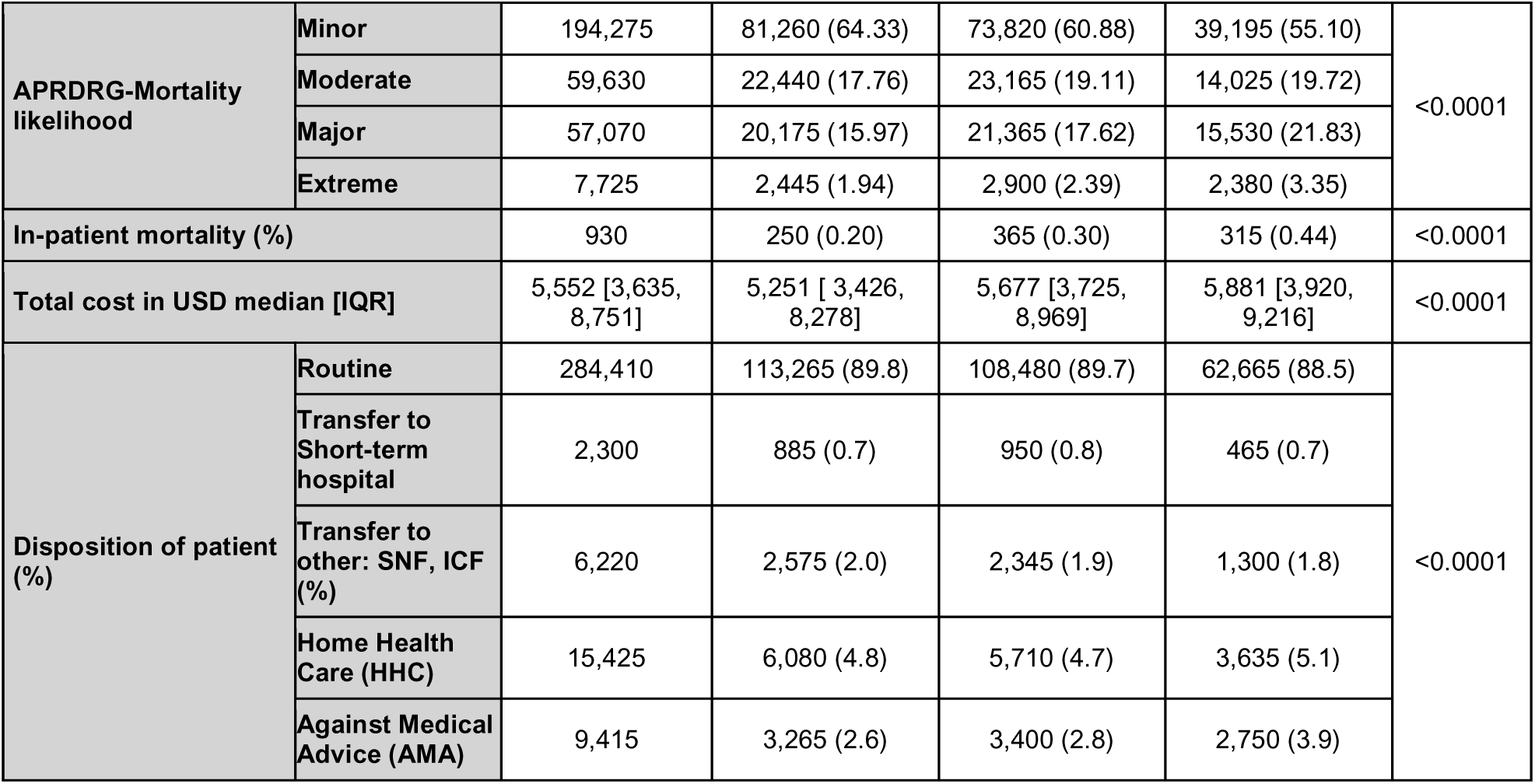
Patient and admission characteristics and outcomes stratified by years.

| Year |  | Overall | 2018 | 2019 | 2020 | P-value |
| --- | --- | --- | --- | --- | --- | --- |
| N (hospital admissions) |  | 63,740 | 25,264 | 24,250 | 14,226 | - |
| Weighted N |  | 318,700 | 126,320 | 121,250 | 71,130 | <0.0001 |
| National Population in the US |  | 986,517,534 | 326,686,501 | 328,329,953 | 331,501,080 | - |
| Rate of Asthma Hospitalizations per 100,000 |  | 32.3 | 38.6 | 37.0 | 21.4 | <0.0001 |
| Demographics |  |  |  |  |  |  |
| Age (median [IQR]) |  | 38 [14,56] | 36 [12,56] | 38 [14,57] | 41 [21,57] | <0.0001 |
| Age Group (%) | 5-19 years | 95,675 | 41,695 (33.0) | 37,115 (30.6) | 16,865 (23.7) | <0.0001 |
|  | 20-44 | 90,890 | 34,100 (27.0) | 34,035 (28.1) | 22,755 (32.0) |  |
|  | 45-64 | 82,545 | 31,400 (24.8) | 31,055 (25.6) | 20,090 (28.2) |  |
|  | 65 and above | 49,590 | 19,125 (15.1) | 19,045 (15.7) | 11,420 (16.1) |  |
| Sex (Female (%)) |  | 202,880 | 79,735 (63.1) | 77,300 (63.8) | 45,845 (64.5) | <0.0001 |
| Race (%) | White | 121,855 | 47,790 (38.9) | 46,085 (38.8) | 27,980 (40.2) | <0.0001 |
|  | Black | 107,490 | 41,860 (34.1) | 41,255 (34.8) | 24,375 (35.0) |  |
|  | Hispanic | 58,115 | 23,430 (19.1) | 22,560 (19.0) | 12,125 (17.4) |  |
|  | Other | 23,805 | 9,860 (8.0) | 8,830 (7.4) | 5,115 (7.4) |  |
| Socio-economic Characteristics |  |  |  |  |  |  |
| Median Household income (%) | 0-25<sup>th</sup> percentile | 117,975 | 46,730 (37.5) | 44,775 (37.4) | 26,470 (37.8) | <0.0001 |
|  | 26<sup>th</sup>-50<sup>th</sup> percentile | 78,370 | 31,595 (25.3) | 28,715 (24.0) | 18,060 (25.8) |  |
|  | 51<sup>st</sup> -75<sup>th</sup> percentile | 67,535 | 26,635 (21.4) | 26,570 (22.2) | 14,330 (20.5) |  |
|  | 76<sup>th</sup> -100<sup>th</sup> percentile | 50,455 | 19,735 (15.8) | 19,580 (16.4) | 11,140 (15.9) |  |
| Urban-rural classes (%) | Metro counties > 1 million | 127,590 | 50,785 (40.4) | 48,465 (40.2) | 28,340 (40.1) | <0.0001 |
|  | Fringe counties of Metro area > 1 million | 77,515 | 30,770 (24.5) | 29,735 (24.6) | 17,010 (24.1) |  |
|  | Counties Metro area of 250000-999,000 | 58,685 | 23,295 (18.5) | 22,300 (18.5) | 13,090 (18.5) |  |
|  | Counties in Metro areas 50,000- 249,999 | 22,645 | 8,845 (7.0) | 8,450 (7.0) | 5,350 (7.6) |  |
|  | Micropolitan areas | 19,020 | 7,385 (5.9) | 7,340 (6.1) | 4,295 (6.1) |  |
|  | Rural areas | 11,700 | 4,775 (3.8) | 4,415 (3.7) | 2,510 (3.6) |  |
| Primary payer (%) | Medicaid | 126,470 | 51,450 (41.63%) | 47,505 (40.19%) | 27,515 (39.70%) | <0.0001 |
|  | Medicare | 68,910 | 26,350 (21.32%) | 26,475 (22.40%) | 16,085 (23.21%) |  |
|  | Private | 88,120 | 35,345 (28.60%) | 33,525 (28.36%) | 19,250 (27.78%) |  |

|  | Self-Pay | 25,735 | 9,690 (7.84%) | 10,010 (8.47%) | 6,035 (8.71%) |  |
| --- | --- | --- | --- | --- | --- | --- |
|  | No Charge | 1,865 | 760 (0.61%) | 690 (0.58%) | 415 (0.60%) |  |
| Comorbidities |  |  |  |  |  |  |
| Obesity (%) |  | 82,710 | 30,880 (24.5) | 31,675 (26.1) | 20,155 (28.3) | <0.0001 |
| Cardiovascular diseases (%) |  | 45,485 | 17,225 (13.6) | 17,120 (14.1) | 11,140 (15.7) | <0.0001 |
| Dyslipidemia (%) |  | 54,195 | 20,245 (16.0) | 20,745 (17.1) | 13,205 (18.6) | <0.0001 |
| Hypertension (%) |  | 113,520 | 43,000 (34.0) | 43,065 (35.5) | 27,455 (38.6) | <0.0001 |
| Diabetes Mellitus (%) |  | 56,570 | 21,980 (17.4) | 21,375 (17.6) | 13,215 (18.6) | <0.0001 |
| Chronic kidney diseases (%) |  | 15,505 | 5,765 (4.6) | 6,020 (5.0) | 3,720 (5.2) | <0.0001 |
| Cancer (%) |  | 3,085 | 1,085 (0.9) | 1,120 (0.9) | 880 (1.2) | <0.0001 |
| Lung Cancer (%) |  | 255 | 110 (0.09) | 90 (0.07) | 55 (0.08) | <0.0001 |
| Allergy and Eczema (%) |  | 32,400 | 13,395 (10.6) | 12,420 (10.2) | 6,585 (9.2) | <0.0001 |
| Chronic sinusitis (%) |  | 7,975 | 3,485 (2.8) | 3,070 (2.5) | 1,420 (2.0) | <0.0001 |
| Sleep apnea (%) |  | 44,435 | 16,810 (13.3) | 17,410 (14.4) | 10,215 (14.4) | <0.0001 |
| GERD (%) |  | 57,825 | 22,130 (17.5) | 22,015 (18.2) | 13,680 (19.2) | <0.0001 |
| Depression (%) |  | 32,930 | 12,340 (9.8) | 12,605 (10.4) | 7,985 (11.2) | <0.0001 |
| Anxiety and related disorders (%) |  | 50,865 | 18,825 (15.0) | 19,005 (15.7) | 13,035 (18.3) | <0.0001 |
| Rheumatoid arthritis and other collagen diseases (%) |  | 17,310 | 6,325 (5.0) | 6,545 (5.4) | 4,440 (6.2) | <0.0001 |
| Liver diseases (%) |  | 4,840 | 1,675 (1.3) | 1,900 (1.6) | 1,265 (1.8) | <0.0001 |
| Substance abuse (%) |  | 24,940 | 9,000 (7.1) | 9,010 (7.4) | 6,930 (9.7) | <0.0001 |
| Characteristics of Hospitalizations |  |  |  |  |  |  |
| Admission quarter (%) | January-March | 100,370 | 36,165 (28.7) | 33,260 (27.5) | 30,945 (43.60) | <0.0001 |
|  | April-June | 72,930 | 30,275 (24.0) | 31,380 (25.9) | 11,275 (15.88) |  |
|  | July-September | 61,735 | 24,920 (19.8) | 24,460 (20.2) | 12,355 (17.41) |  |
|  | October-December | 82,995 | 34,710 (27.5) | 31,880 (26.4) | 16,405 (23.11) |  |
| LOS in days median [IQR] |  | 2 [1,3] | 2 [1,3] | 2 [1,3] | 2 [1,4] | <0.0001 |
| LOS >7 days |  | 19,055 | 7,595 (6.0) | 7,100 (6.0) | 4,360 (6.1) | <0.0001 |
| APRDRG-Asthma severity (degree of loss of function) | Minor | 121,590 | 45,495 (36.02) | 50,535 (41.7) | 25,560 (35.9) | <0.0001 |
|  | Moderate | 108,575 | 45,915 (36.35) | 39,235 (32.4) | 23,425 (32.9) |  |
|  | Major | 73,230 | 27,630 (21.87) | 26,885 (22.2) | 18,715 (26.3) |  |
|  | Extreme | 15,305 | 7,280 (5.76) | 4,595 (3.8) | 3,430 (4.8) |  |
| **APRDRG-Mortality likelihood** | **Minor** | 194,275 | 81,260 (64.33) | 73,820 (60.88) | 39,195 (55.10) | <0.0001 |
|  | **Moderate** | 59,630 | 22,440 (17.76) | 23,165 (19.11) | 14,025 (19.72) |  |
|  | **Major** | 57,070 | 20,175 (15.97) | 21,365 (17.62) | 15,530 (21.83) |  |
|  | **Extreme** | 7,725 | 2,445 (1.94) | 2,900 (2.39) | 2,380 (3.35) |  |
| **In-patient mortality (%)** |  | 930 | 250 (0.20) | 365 (0.30) | 315 (0.44) | <0.0001 |
| **Total cost in USD median [IQR]** |  | 5,552 [3,635, 8,751] | 5,251 [ 3,426, 8,278] | 5,677 [3,725, 8,969] | 5,881 [3,920, 9,216] | <0.0001 |
| **Disposition of patient (%)** | **Routine** | 284,410 | 113,265 (89.8) | 108,480 (89.7) | 62,665 (88.5) | <0.0001 |
|  | **Transfer to Short-term hospital** | 2,300 | 885 (0.7) | 950 (0.8) | 465 (0.7) |  |
|  | **Transfer to other: SNF, ICF (%)** | 6,220 | 2,575 (2.0) | 2,345 (1.9) | 1,300 (1.8) |  |
|  | **Home Health Care (HHC)** | 15,425 | 6,080 (4.8) | 5,710 (4.7) | 3,635 (5.1) |  |
|  | **Against Medical Advice (AMA)** | 9,415 | 3,265 (2.6) | 3,400 (2.8) | 2,750 (3.9) |  |

The cost of an asthma admission surged across all geographic divisions in 2020 compared to the pre-pandemic period (P<0.05). Notably, the Pacific division had the highest hospitalization costs compared to other divisions annually between 2018 and 2020 (P<0.05) (Figure 2). Over the span of three years, there was a notable increase in hospitalization costs within this division (P=0.004), which was particularly evident when comparing 2020 to 2018 (P=0.005), although the increase between 2019 and 2020 did not reach statistical significance.

**Figure 2:**
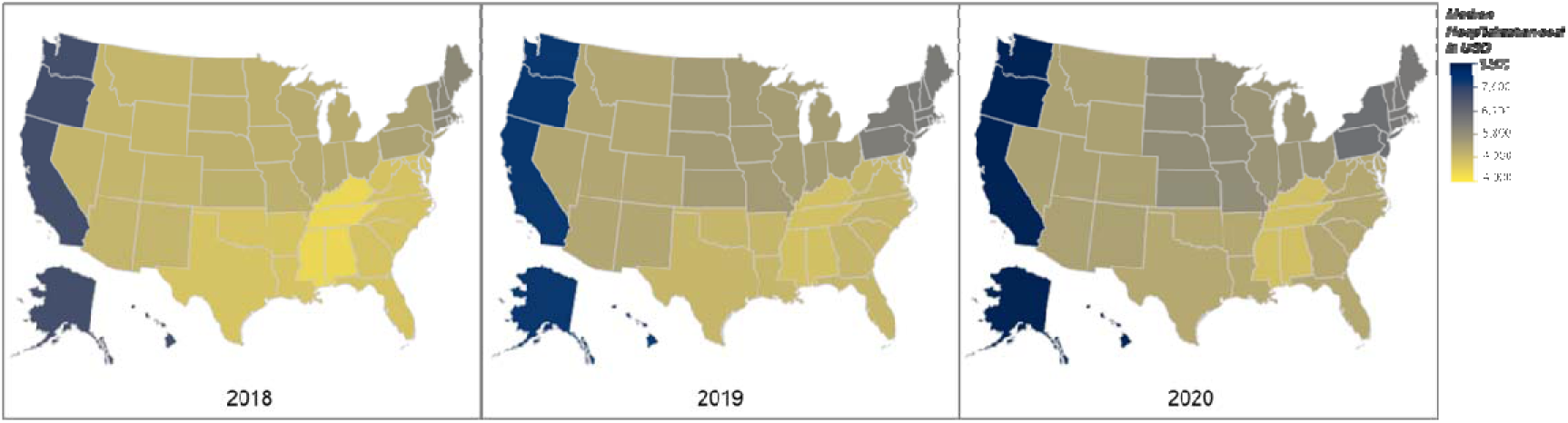
Census division variation in the median cost of asthma hospitalization during the pandemic (2020) compared to pre-pandemic (2019 and 2018). The cost of asthma hospitalizations in every division significantly increased in 2020 compared to pre-pandemic (P < 0.05). The Pacific division had the highest hospitalization costs compared to other divisions each year (P < 0.05); There is also a significant increase in hospitalization cost in this division in 2020 compared to 2018 (p=0.005) but no significant increase between 2020 and 2019 (p>0.05).

Using a multivariable linear regression model adjusted for patients’ demographic characteristics, socioeconomic status, and chronic medical comorbidities, we found the cost of asthma admission in 2020 was 6.2% higher than pre-pandemic period, corresponding to an adjusted exponential of the slope for cost (95% CI) of 1.03 (1.03-1.04) (Table 2). Figure E2 provides a more detailed stratification of yearly asthma hospitalization cost by quarter.

**Table 2:** Univariate and multivariable regression analysis of study outcomes, adjusted for demographics, comorbidities, and hospital admission characteristics.

|  | COVID-19 period |  | Pre-COVID-19 period |  |
| --- | --- | --- | --- | --- |
|  | OR (95% CI) | aOR (95%) | OR (95% CI) | aOR (95%) |
| *In-patient mortality* | 1.79 (1.31-2.42) | 1.27 (1.10-1.50) | 0.56 (0.41-0.76) | 0.78 (0.67-0.92) |
| *LOS≥7 days* | 1.03 (1.01-1.07) | 0.95 (0.91-0.98) | 0.97 (0.93-0.99) | 1.05 (1.01-1.09) |
|  | Exponential of slope (95% CI)* | Adjusted Exponential of slope (95% CI)<sup>§</sup> | Exponential of slope (95% CI) | Adjusted Exponential of slope (95% CI) |
| *Cost* | 1.04 (1.03-1.05) | 1.03 (1.03-1.04) | 0.96 (0.95-0.97) | 0.97 (0.96-0.97) |
| *LOS* | 1.01 (1.00-1.01) | 0.99 (0.98-0.99) | 0.99 (0.99-1.00) | 1.01 (1.01-1.02) |
| Percentage Change in cost= $\frac{e^{COVID-19} - e^{Pre-COVID-19 period}}{e^{Pre-COVID-19 period}} \times 100$ | | | | |
| *Unadjusted percent change =+8.3% |  |  |  |  |
| § adjusted percent change =+6.2% |  |  |  |  |

### Annual trends in asthma hospitalizations

Overall, 63,740 weighted asthma admissions were recorded between 2018 and 2020 (Table 1). The rate of asthma hospitalization decreased from 38.6 per 100,000 in 2018 to 21.4 per 100,000 in 2020 (P <0.0001), with the most notable decline between April and June (Figure 1-B, E*5*).

The overall median age of patients admitted for asthma was 38 [14,56] years. There was a significant increase in admissions among patients aged 20-44 and 45-64 years (P<0.0001) over the 3 years. Females demonstrated a modest but significant rise in hospitalizations from 63.1% in 2018 to 64.5% in 2020 (P<0.0001). Additionally, there was an increasing trend in asthma admissions for White and Black patients, while hospitalizations for Hispanic patients decreased (Table 1).

Comorbidities such as obesity (24.5% in 2018 to 28.3% in 2020), cardiovascular diseases (13.6% in 2018 to 15.7% in 2020), and anxiety (15.0% in 2018 to 18.3% in 2020) saw slight increases, while substance abuse rose notably in 2020 (7.1% in 2018 to 9.7% in 2020) (P <0.0001) (Table 1).

The All Patient Refined Diagnosis Related Groups (APRDRG) are a clinical model that classifies patients based on their severity of illness, risk of mortality and reason for admission. The APRDRG of illness showed a slight increase in cases classified as major (21.87% in 2020) and extreme (5.76% in 2020). The risk of mortality categories reflected higher severity, with extreme likelihood of mortality rising to 3.35% in 2020 from 2.39% in 2019 and 1.94% in 2018. Most patients were discharged routinely (89.8% in 2020), with slight increases in home health care transfers (5.1%) and discharges against medical advice (3.9%) in 2020 (Table 1).

Asthma hospitalization rates decreased in all divisions in 2020 compared to pre-pandemic levels (P <0.0001) (Figure 3). The Mid-Atlantic division consistently had the highest hospitalization rates each year (P <0.001). This division experienced a significant reduction in hospitalizations over the three years (P<0.01), with marked decreases in 2020 compared to both 2018 (P <0.04) and 2019 (P<0.04).

**Figure 3:**
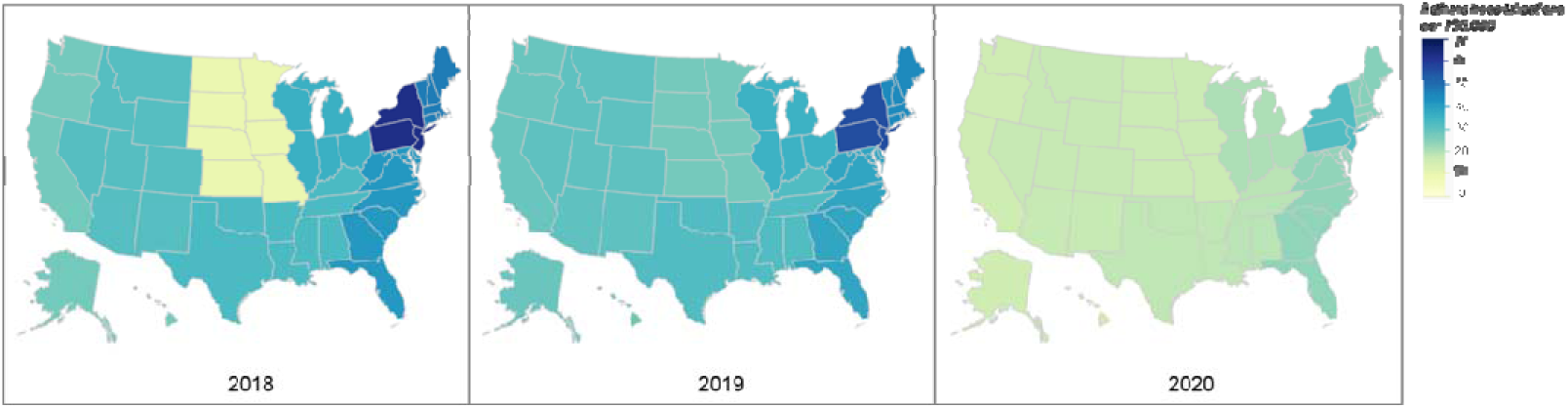
Census division variation in the rate of asthma hospitalization per 100,000 during the pandemic (2020) compared to pre-pandemic (2019 and 2018). The rate of asthma hospitalizations in every division significantly decreased in 2020 compared to pre-pandemic (P < 0.0001). The Mid-Atlantic division had the highest hospitalization rates compared to other divisions each year (P < 0.001); there was also a significant decrease in hospitalizations in this division over the 3 years (p=0.01) and in 2020 compared to 2018 (p=0.04) and 2019 (p=0.04).

Further stratification of annual asthma hospitalization characteristics by quarter are detailed in figure E6 and E7.

### Annual trends in hospital mortality

In-patient mortality rates modestly increased in 2020 (0.44%) compared to 2019 (0.30%) and 2018 (0.20%) (Table 1). During the pandemic, Q3 had the lowest mortality rate (Figure 1-C). In 2018, the East North Central division had the highest mortality rate among all divisions (P<0.05), whereas in 2019, the Mountain division recorded the highest rate (P<0.05). During the pandemic, the Mid-Atlantic division experienced the highest mortality rate compared to other divisions (P<0.05) (Figure 4).

**Figure 4:**
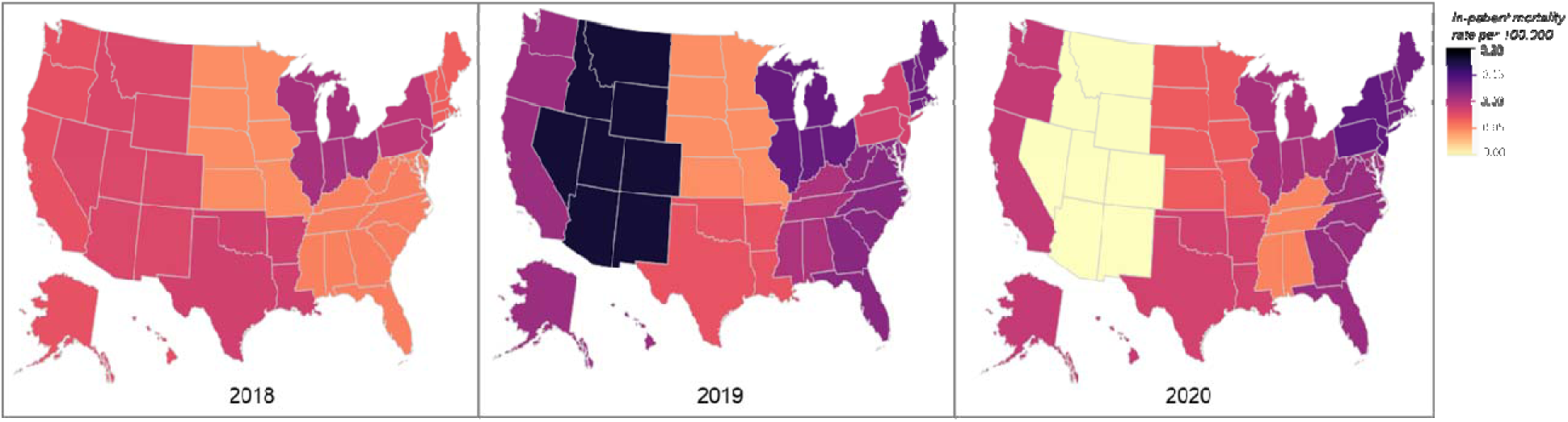
Census division variation in the in-patient asthma mortality rate per 100,000 during the pandemic (2020) compared to pre-pandemic (2019 and 2018). Compared to other divisions, the East North Central division had the highest mortality rate in 2018 (P < 0.05), while in 2019 the Mountain division had the highest mortality rate (P < 0.05). During the pandemic, the Mid-Atlantic division had the highest mortality rate compared to other divisions (P < 0.05).

Using a multivariable linear regression model adjusted for patients’ demographic characteristics, socioeconomic status, and chronic medical comorbidities, our analysis revealed odds of in-patient mortality were 27% higher during the COVID-19 pandemic than in the pre-pandemic period (aOR (95% CI): 1.27 (1.10-1.50)) (Table 2).

Figure E9 provides a more detailed stratification of yearly in-patient mortality by quarter.

### Annual trends in length of stay

The overall median LOS was 2 [1,3] days, which remained consistent across the 3 years and the percentage of patients with a LOS >7 days remained stable at 6.0% (Table 1). Q2 had the lowest proportion of patients with LOS >7 days (Figure 1, Figure E8). During early pandemic, admission with LOS >7 days decreased in all divisions compared to pre-pandemic levels, except for the Mid-Atlantic, East North Central, and Mountain divisions. The Mid-Atlantic division consistently had the highest rate of hospitalizations with LOS>7 days compared to other divisions in every year (p <0.05) (Figure 5).

**Figure 5:**
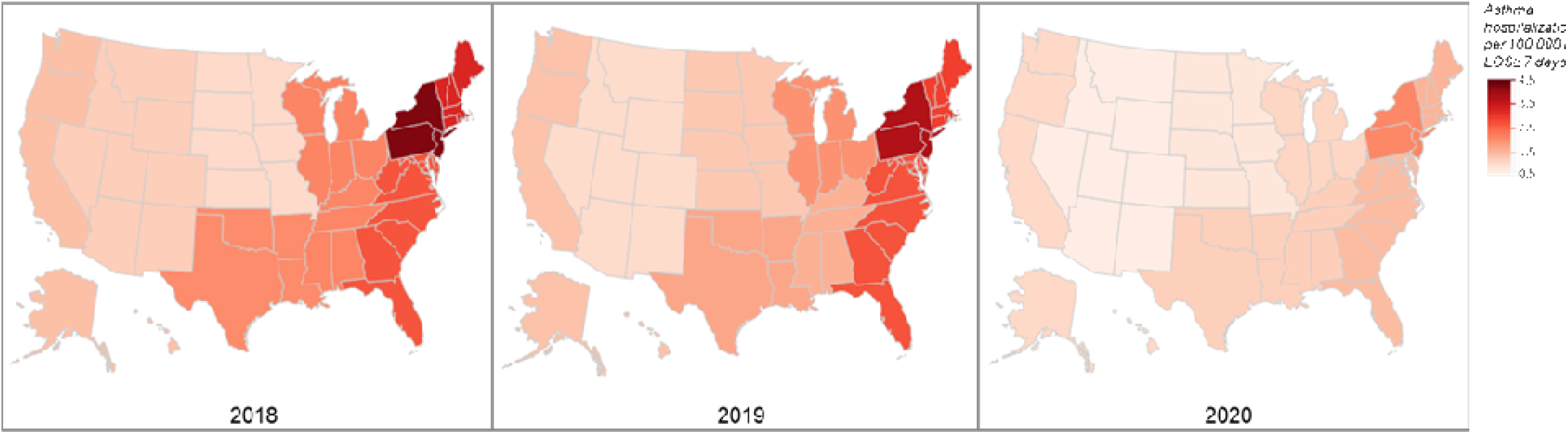
Census division variation in the percentage of inpatient length of stay (LOS) longer than 7 days during the pandemic (2020) compared to pre-pandemic (2019 and 2018). Except for the Mid-Atlantic, East North Central, and Mountain divisions (p>0.05), the percentage of admissions with LOS> 7 days significantly decreased in all other divisions in 2020 compared to pre-pandemic (P < 0.05). The Mid-Atlantic division had the highest rate of hospitalization with LOS>7 days compared to other divisions every year (P < 0.05).

Using a multivariable linear regression model adjusted for patients’ demographic characteristics, socioeconomic status, and chronic medical comorbidities, we found that patients with asthma were more likely to have a shorter admission (exponential of slope for LOS (95% CI) during the COVID-19 period: 0.99 (0.98-0.99)). In addition, the odds of asthma admission of more than 7 days were 5% lower during COVID-19 pandemic than in the pre-pandemic period (aOR (95% CI): 0.95 (0.91-0.98)) (Table 2).

Figure E9 provides a more detailed stratification of yearly LOS by quarter.

### COVID-19 hospitalizations with secondary diagnosis of asthma

To differentiate healthcare utilization patterns between COVID-19 and asthma admissions, we conducted a secondary analysis on the NIS cohort of 1,079,310 individuals with a primary diagnosis of COVID-19 and a secondary diagnosis of asthma. The admission rate reached 325.6 per 100,000. We found that 508,910 (47%) of the patients were female, with a median age of 66 years [53,77]. Additionally, 547,295 (50%) were admitted during the last quarter of the year, with a median length of stay (LOS) of 5 days [3,9]. The South Atlantic division had the highest admission rate compared to other geographic divisions, accounting for 218,890 (20%) of the total admissions (Figure E10).

## Discussion

### Main findings

The start of the COVID-19 pandemic was marked by a shift in healthcare utilization and economic burden. Using the NIS-HCUP dataset, we found that the median cost of asthma hospitalizations rose from $5,251 in 2018 to $5,881 in 2020, with significant geographic disparities, particularly in the Pacific division. Despite the rising costs, the rate of asthma hospitalizations significantly dropped from 38.6 per 100,000 in 2018 to 21.4 per 100,000 in 2020. The demographics of admitted patients revealed an increase in severity and comorbidities, such as obesity and cardiovascular diseases, while in-patient mortality rates modestly increased during the pandemic. However, the length of stay remained stable. In comparison, COVID-19 hospitalizations with a secondary diagnosis of asthma indicated significant healthcare utilization, with the South Atlantic region having the highest admission rates. Though the rate of hospitalizations for asthma as a primary diagnosis decreased, the rate of hospitalizations for asthma as a secondary diagnosis to COVID-19 increased.

### Cost of asthma hospitalizations

According to an analysis of the Medical Expenditure Panel Survey asthma imposes a significant and increasing economic burden on the US^5^, with an estimated $960 billion in direct and indirect costs projected between 2019 and 2038^25^. In 2013, the annual per-person cost of hospitalization was $529^5^. In 2020, the COVID-19 pandemic aggravated this burden, with a notable increase in admission costs by more than $700 from 2018, resulting in a median cost of $5,881. We noted a 6.2% rise in asthma hospitalization costs which is higher than the general inflation rate for medical services of 2.8% in 2020^20^. We speculate that this increase in asthma admission costs could be attributed to several reasons, including loss of revenue by US hospitals. The American Hospital Association revealed that US hospitals experienced a total loss of over $200 billion between March 1 and June 30, 2020. This was primarily due to an estimated 45% decrease in operating revenue driven by a reduction in elective procedures and routine care as hospitals focused on treating COVID-19 patients^26^. Second, during the pandemic there has been an increase in operating expenses, driven by shortages and unprecedented inflation that escalated the costs of medical supplies, drugs, and equipment^20,27,28^. It was estimated that drug expenses increased by 36.9% and medical supplies by 0.6% from pre-pandemic period^27^. Third, a significant workforce shortage has emerged as caregivers have been affected by COVID-19, forcing hospitals to rely heavily on more expensive contract labor and increase hazard pay^27,28^. Fourth, there has been an increase in patient acuity as the result of delaying or deferring care caused by fears of related to COVID-19^29,3029,31^. We found that admitted patients showed higher degrees of loss of function due to asthma, had more comorbidities, and experienced prolonged lengths of stay. Consequently, this resulted in higher healthcare expenditures and increased costs.

We found that highest hospitalization costs were in the Pacific division of the US. This variation in hospitalization costs across different regions of the US aligns with findings from previous studies^32^ which showed regional disparities in healthcare expenses. This could be attributed to differences in local healthcare policies, the cost of living, patient population characteristics, and hospital operational expenses.

### Healthcare Utilization patterns

The early phase of the COVID-19 pandemic saw a significant decrease in asthma admissions^8,11,15^ compared to pre-pandemic levels. The most pronounced decrease in asthma admissions occurred between April and June 2020, coinciding with the initial phases of the COVID-19 pandemic when lockdowns and preventative measures were most stringent. This decline in asthma admissions could be due to several factors. Lockdown measures reduced exposure to common asthma triggers such as outdoor pollutants and respiratory infections, possibly leading to fewer exacerbations^33^. Behavioral changes included adherence to medications and preventive measures such as wearing masks and social distancing^16,34,35^. Additionally, concerns about contracting COVID-19 in healthcare settings discouraged asthma patients from seeking emergency care unless absolutely necessary^36^. The observed decline in asthma admissions during COVID-19 underscores the impact of public health measures and patient behavior on disease outcomes.

Despite a national decline in asthma hospitalization rates between 2018 and 2020, the Mid-Atlantic division, which includes New York, New Jersey, and Pennsylvania, had higher rates of asthma admissions. Although air quality improved during lockdowns^33^, individuals with asthma in the Mid-Atlantic might have been more exposed to poor indoor air quality, or to indoor allergens such as dust mites, mold, pet dander^39^. Another possible reason could be disparities in access to care, which could lead to higher hospitalization rates^39^.

### In-patient mortality during asthma hospitalization

We observed an increase in mortality rate, with a 27% higher likelihood of death during the pandemic compared to the pre-pandemic period^40,41^.The sudden surge in COVID-19 cases overwhelmed healthcare facilities^7,41^. Many individuals requiring routine medical attention experienced cancellations and delays in receiving treatment^30^. These disruptions increased the likelihood of disease progression, elevating the risk of severe illness and mortality among non-COVID-19 patients^41^. Moreover, patients admitted to hospitals were often older and from lower socio-economic backgrounds^40^. Many patients had pre-existing chronic conditions, which compounded their vulnerability^40^. The convergence of these factors led to an increase in patient mortality as the healthcare system struggled to provide adequate care amidst the unprecedented demand^41^. The Mid-Atlantic division recorded the highest mortality rate compared to other divisions.

### Length of stay (LOS) of asthma hospitalizations

The LOS remained consistent across the three years with a 5% reduced likelihood of being admitted for more than 7 days. This finding differs from that reported by McAlister et al., where patients hospitalized with non-COVID-19 conditions had a mean LOS of 9 days both before and during the pandemic^42^. Another study reported a longer hospital stay among non-COVID-19 patients during the same period^31^. A reason for this discrepancy could be variations in healthcare system capacity, patient demographics, or local pandemic management strategies that influenced hospital workflows and resource availability. Those factors could account for the observed geographic division disparities in LOS in our study. The lowest proportion of patients with prolonged LOS was reported in Q2. The surge of COVID-19 cases in Q2 in March 2020 led to hospitals reaching or exceeding capacity. Hospitals allocated nearly all their resources to treating COVID-19 patients, leaving a little room for non-COVID-19 cases such as asthma^30^.

### COVID-19 vs. asthma admissions during the pandemic

When compared to asthma admissions, the rate of hospitalization for COVID-19 with a secondary asthma diagnosis was 15 times higher. In contrast to the demographics of asthma hospitalizations, males with chronic asthma were more frequently admitted for COVID-19, consistent with existing literature. In addition, patients with COVID-19 were older and stayed longer in the hospital due to more severe disease in the setting of their chronic airway inflammation and underlying comorbidities^44,45^. Furthermore, we observed the highest rate of COVID-19 in individuals with asthma in the South Atlantic division.

During the early pandemic period, healthcare utilization patterns differed between patients with asthma hospitalized for COVID-19 and those admitted solely for asthma as COVID-19 admissions required greater healthcare resources.

### Limitations

A major limitation of this study is the use of asthma as a primary diagnosis. Over one million individuals were admitted to the hospital with COVID-19 as a primary diagnosis and asthma as a secondary diagnosis in 2020, thus most individuals with respiratory conditions were more likely to have COVID-19 as a primary diagnosis and asthma as a secondary diagnosis. Therefore, the contribution of asthma to the hospitalizations, mortality, and length of stay in our study is likely underestimated. A second limitation is its observational approach, thus we are unable to establish causal effect. Despite these limitations, the HCUP-NIS data offers many advantages in asthma research in that it is a large nationally representative sample drawn from patient health records.

## Conclusion

We found that the COVID-19 pandemic was associated with decreased asthma hospitalizations and increased healthcare costs. The social distancing protocols and policies during the pandemic may have reduced asthma exacerbations requiring admission. However, avoidance of the healthcare system may have increased asthma severity and healthcare cost for those who were admitted. Our findings suggest that pandemic mitigation strategies are needed to educate patients with asthma on when to seek medical care to avoid adverse outcomes.

## Data Availability

All data produced in the present study are available upon reasonable request to the authors.

## Acknowledgements

Dr. Serpil Erzurum

## Funding/support

Cleveland Clinic Research Program Committee Grant

## Financial/nonfinancial disclosures

None declared.

## Author Contributions

**Neha Solanki:** Conceptualization, methodology, software, formal analysis, writing-review and editing, project administration; **Battoul Fakhry:** methodology, software, formal analysis, investigation, data curation, writing-original draft; **Peng Zhang:** formal analysis, writing-review and editing**; Amy Attaway:** formal analysis, writing-review and editing**; Joe Zein:** resources, writing-review and editing **; Wayne Tsuang:** writing-review and editing, visualization, supervision

## Conflict of Interest

The authors have no relevant conflicts of interest to disclose.

## Ethics

Our paper does not require ethical approval. The data from NIS are de-identified, thus exempting review

## Abbreviations

AHRQ: Agency for Healthcare Research and Quality
APR-DRG: All Patient Refined Diagnosis Related Group
CCR: Cost-To-Charge Ratios
COPD: Chronic Obstructive Pulmonary Disease
ED: Emergency Department
HCUP: Healthcare Cost and Utilization Project
NIS: National Inpatient Sample
US: United States
USD: United States Dollars

## Notes

### Competing Interest Statement

The authors have declared no competing interest.

### Funding Statement

The study was funded by Cleveland Clinic Research Program Committee.

### Author Declarations

The data is found at HCUP-NIS: https://hcup-us.ahrq.gov/nisoverview.jsp.

