## Supplementary figures and images for "Direct Health Care Costs Associated with Asthma Hospitalizations Before and During the Covid-19 Pandemic in the United States: A Nationwide Inpatient Sample Analysis"

### Supplemental File

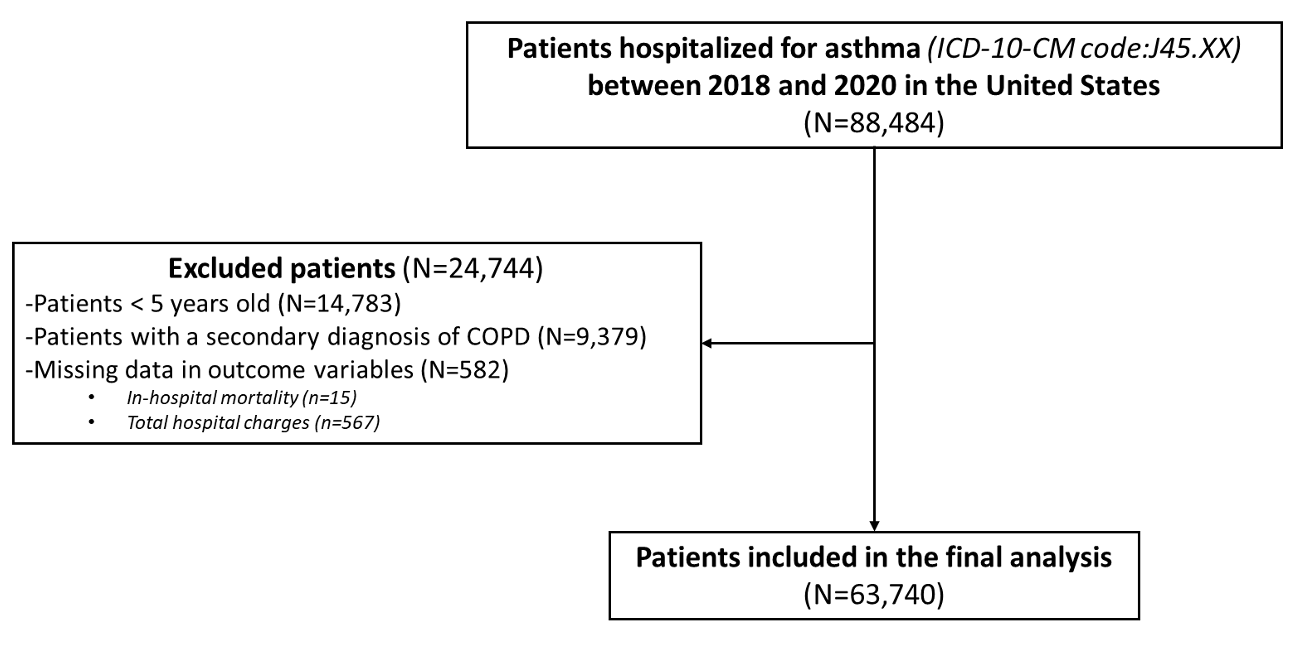
Figure Legend.

Figure E1: Flowchart of patient selection.

Figure E1.
